# The Age-Related Probability of Dying from COVID-19 among Those Infected: A Relative Survival Analysis

**DOI:** 10.1101/2022.01.26.22269928

**Authors:** David A. Swanson, Dudley Poston, Steven G. Krantz, Arni S.R. Srinivasa Rao

**Affiliations:** Department of Sociology, University of California Riverside, Riverside, CA USA 9252; Center for Studies in Demography & Ecology, University of Washington, Seattle, WA USA 98195; Department of Sociology, Texas A & M University, College Station, TX USA 77843; Department of Mathematics, Washington University in St. Louis, Campus Box 1146, One Brookings Drive, St. Louis, Missouri 63130 USA; Laboratory for Theory and Mathematical Modeling, Department of Medicine-Division of Infectious Diseases, Medical College of Georgia, 1120 15th Street, AE2040, Augusta, GA 30912, U.S.A.

**Keywords:** China, confidence interval, Gompertz Model, Hubei Province, life table, superpopulation, Wuhan

## Abstract

**Background:** COVID-19 was first identified in Wuhan, the capital city of the province of Hubei in China. Due to the presentation of multiple symptoms at the same time, it is clinically important to understand the probability of dying from COVID-19 vs. the probability of dying from other causes.

**Methods:** Using data collected in Hubei that identified by age those who died of COVID-19 or its sequelae among the infected, we constructed a life table showing the conditional probability of dying at age x from COVID-19 and its sequela among those infected. Following the relative survival perspective, we also computed corresponding data for China that matched the format of the life table we constructed from the Hubei study. We then formed ratios of the 10-year conditional portability of dying at age x from COVID-19 for the Hubei COVID-19 victims to the ten-year conditional probability of dying at age x from all non-COVID-19 causes for those not infected by COVID-19 in China as a whole.

**Findings:** At every age, the conditional probability of dying from COVID-19 among those infected in Hubei is higher than the conditional probability of dying from all non-COVID-19 causes for China as a whole. Following a general age-related mortality pattern, the conditional probability of dying from COVID-19 from age 20 onward increases monotonically for those who are infected. Relative to the probability of dying in China from all other causes for those not infected, however, it declines monotonically from age 20 to age 70.

**Interpretation:** At younger ages the relative conditional probability of dying from CVOD-19 among the infected is substantially higher than it is for those infected who dying of all other causes and while staying higher at all ages, it declines monotonically with age. The monotonic decline in the ratio from age 20 to age 70 is a result of the age-related increase in the probability of dying from one or more of a number of competing causes, which, in the case at hand is manifested in the fact that non-COVID-19 deaths in China among the uninfected were generally increasing at a faster age-related rate than were the COVID-19 deaths to the infected in Hubei.

## Introduction

As of January 19^th^, 2022 the total worldwide COVID-19 related deaths from the beginning of the pandemic in 2019 are around 5.55 million and total infections are around 334 million as of January 19^th^, 2022 (Ritchie et al., 2022). Some researchers have measured the impact of these excess deaths on population life expectancy, while others argued that life expectancy is reduced due to COVID-19 deaths (Trias-Llimós and Bilal 2020, Andrasfay and Goldman 2021, Castro et al., 2021, Islam et al., 2021; Venkataramani, O’Brien and Tsai 2021; Woolf, 2021). In general, the approach considered in these studies is one of the commonly accepted techniques using period life tables (Chan, Cheng, and Martin 2021, Yusuf, Martins, and Swanson 2014). However, the middle and older-aged excess deaths during 2020-21 influence the computation of life expectancy of newborn babies of 2020-21 in all of them. In essence, these studies assume that the current mortality rates among adults will have the same pattern when a current newborn reaches adulthood, and, accordingly, survival probabilities were computed. However, a newborn in 2020 need not be influenced by COVID-19 excess adult deaths during 2020-21 because these deaths may not occur every year for the next 20-30 years (Rao, Swanson, Krantz, 2022). This problem can be avoided in several ways. One is to examine Person-years of Life Lost (Andersen, Canudas-Romo and Keiding, 2013; Pifarré i Arolas et al. 2021, Swanson et al. 2022), another is to compute the loss of expected remaining life (Goldstein and Lee 2020, Heuveline 2021, Rao, Swanson, Krantz (2022)). A third approach is to compare the probability of dying by age of those who have contracted COVID-19 to those who have not been subject to the COVID-10 pandemic, a “relative survival” technique (Stroup et al., 2014). In this paper we take this latter approach in the form of a case study of COVID-19 victims in Hubei, China.

## Background

Hubei is a province in China of 72,400 square miles and a population in excess of 57 million (Britannica, no date). COVID-19 symptoms such as viral sore throat, cough, pneumonia, fever, myalgia and fatigue were first clinically seen among hospitalized patients in its capital, Wuhan, during December 2019 (Chen et al., 2020). The first clinical studies were conducted at hospitals in Wuhan after ruling out common bacterial and viral pathogens that cause pneumonia (Huang et al. (2020). Clinical information such as chest images, complete blood count, sputum tests, coagulation profile, and liver tests were conducted. Due to multiple symptoms presenting at the same time, understanding the relative survival of diagnosed COVID-19 patients became clinically important (Stroup et al., 2014). In January 2020, the SARS-CoV-2 virus was officially confirmed and quarantine zones were implemented across the province, especially in the city of Wuhan, which was identified as the epicenter of outbreak (Verity et al. 2020). Given this history, it is not surprising that one of the first collections of death data by age for COVID-19 victims was collected in Hubei Province, a subject to which we now turn.

## Data and Methods

In this study we employ the COVID-19 death data by age for Hubei Province taken from Table 1 in Verity et al. (2020). The study population was confirmed to be COVID-19 positive and the deaths are directly due to COVID-19 and its immediate sequelae. The death and population data were collected such that each subject had less than one year of exposure. No adjustment was made for this so the assumption is that each subject had one year of exposure. The data are subject to right censoring and other issues. For details see Verity et al. (2020). The data are presented in a manner consistent with a clinical study, which along with the sample size (n= 44,672, with 1,023 recorded deaths by age) makes it feasible to construct a life table (Swanson, 2021; Swanson, Chow, and Bryan, 2020), a rarity so far among COVID-19 studies.

**Table 1.**
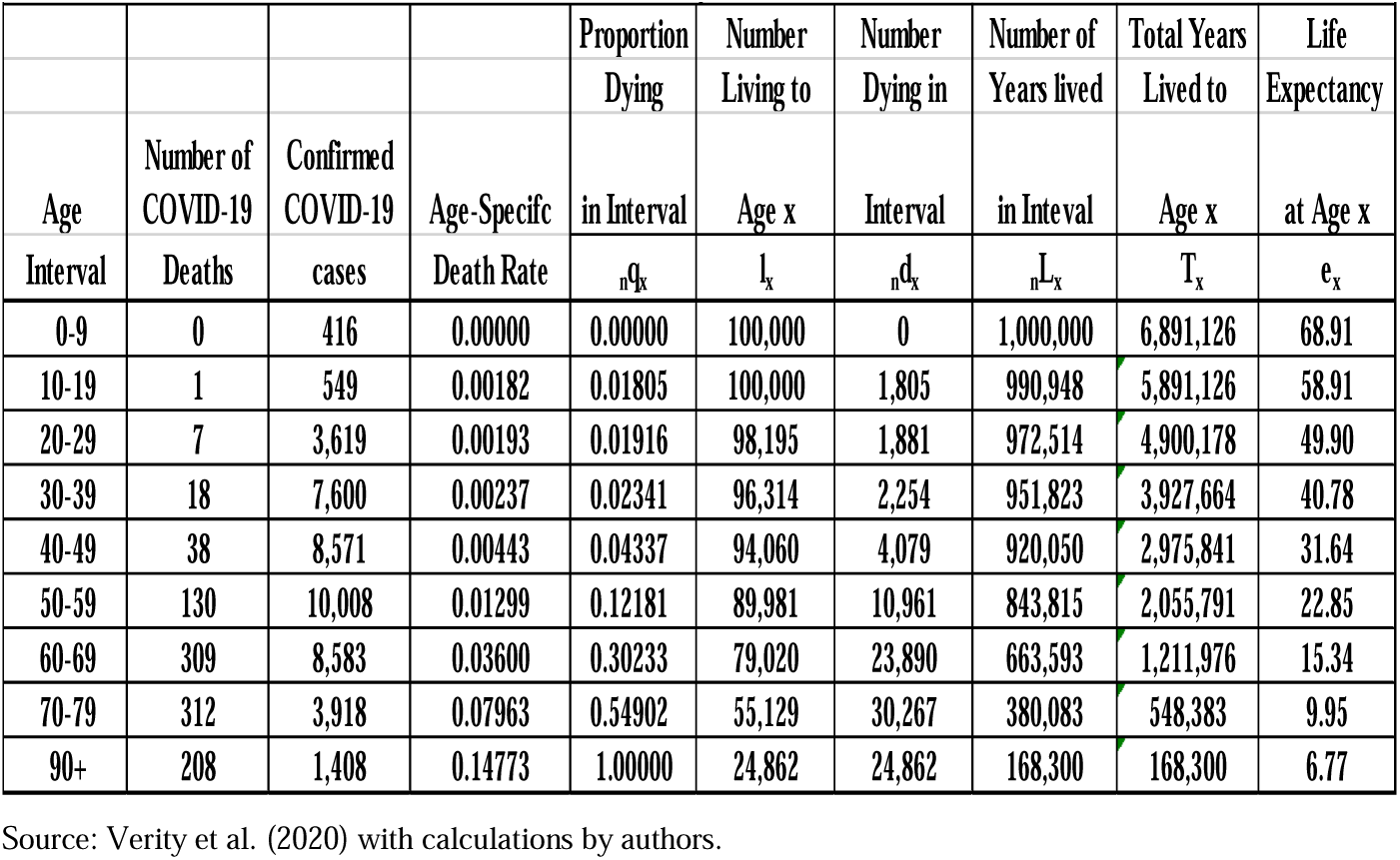
Life Table for COVID-19 Sample from Hubei, China.

| Table 1. Life Table for COVID-19 Sample from Hubei, China |  |  |  |  |  |  |  |  |  |
| --- | --- | --- | --- | --- | --- | --- | --- | --- | --- |
|  |  |  |  | Proportion | Number | Number | Number of | Total Years | Life |
|  |  |  |  | Dying | Living to | Dying in | Years lived | Lived to | Expectancy |
| Age | Number of | Confirmed | Age-Specific |  |  |  |  |  |  |
| Interval | Deaths | cases | Death Rate | in Interval | Age x | Interval | in Interval | Age x | at Age x |
| | | | | ${}_nq_x$ | $l_x$ | ${}_nd_x$ | ${}_nL_x$ | $T_x$ | $e_x$ |
| 0-9 | 0 | 416 | 0.00000 | 0.00000 | 100,000 | 0 | 1,000,000 | 6,891,126 | 68.91 |
| 10-19 | 1 | 549 | 0.00182 | 0.01805 | 100,000 | 1,805 | 990,948 | 5,891,126 | 58.91 |
| 20-29 | 7 | 3,619 | 0.00193 | 0.01916 | 98,195 | 1,881 | 972,514 | 4,900,178 | 49.90 |
| 30-39 | 18 | 7,600 | 0.00237 | 0.02341 | 96,314 | 2,254 | 951,823 | 3,927,664 | 40.78 |
| 40-49 | 38 | 8,571 | 0.00443 | 0.04337 | 94,060 | 4,079 | 920,050 | 2,975,841 | 31.64 |
| 50-59 | 130 | 10,008 | 0.01299 | 0.12181 | 89,981 | 10,961 | 843,815 | 2,055,791 | 22.85 |
| 60-69 | 309 | 8,583 | 0.03600 | 0.30233 | 79,020 | 23,890 | 663,593 | 1,211,976 | 15.34 |
| 70-79 | 312 | 3,918 | 0.07963 | 0.54902 | 55,129 | 30,267 | 380,083 | 548,383 | 9.95 |
| 90+ | 208 | 1,408 | 0.14773 | 1.00000 | 24,862 | 24,862 | 168,300 | 168,300 | 6.77 |

Source: Verity et al. (2020) with calculations by authors.

From these data, the life table shown as Table 1 was constructed using Fergany’s Method (Fergany 1971). As can be seen in Table 1. Fergany’s (1971) method is advantageous because only the age-specific death rates are needed to construct an abridged life table. “In addition to its simplicity, it is, in contrast to other methods, self-contained in the sense that beyond making only the assumption of approximating the force of mortality by a step function (which is all we observe anyway) no further assumptions, approximations, or parameter estimates are required to compute all the life table functions.” (Fergany 1971: 334).

From a 2016 abridged life table for China (Appendix Table 1) that was by gender (National Bureau of Statistics of China, no date), we computed the weighted number of survivors at age x (l_x_) for both genders combined (Kintner, 2004: 318-319) using the 2020 population age distribution by gender taken from the National Bureau of Statistics of China as provided by Statistica (no date). We then found the l_x_ values at 20, 30,…70, and 80 for both genders combined (also known as “all sexes”) and computed the conditional probabilities of dying (_10_q_x_) in the next ten year period given that one had lived to age x. These are shown in Table 1. Because everybody who reached age 80 dies in the subsequent “open age” interval, the probability of dying is set to 1.00 at age 80.

The appendix shows the mathematics underlying the conditional probability of dying and surviving from one age to the next.

## Results

Starting at age 20, Table 2 shows the _10_q_x_ values for the Hubei COVID-19 victims in 2020 and the _10_q_x_ values derived from the 2016 life table for China. Figure 1 displays these same results in graphic form. Figure 3 shows the ratio of these same _10_q_x_ values in graphic form.

**Table 2.**
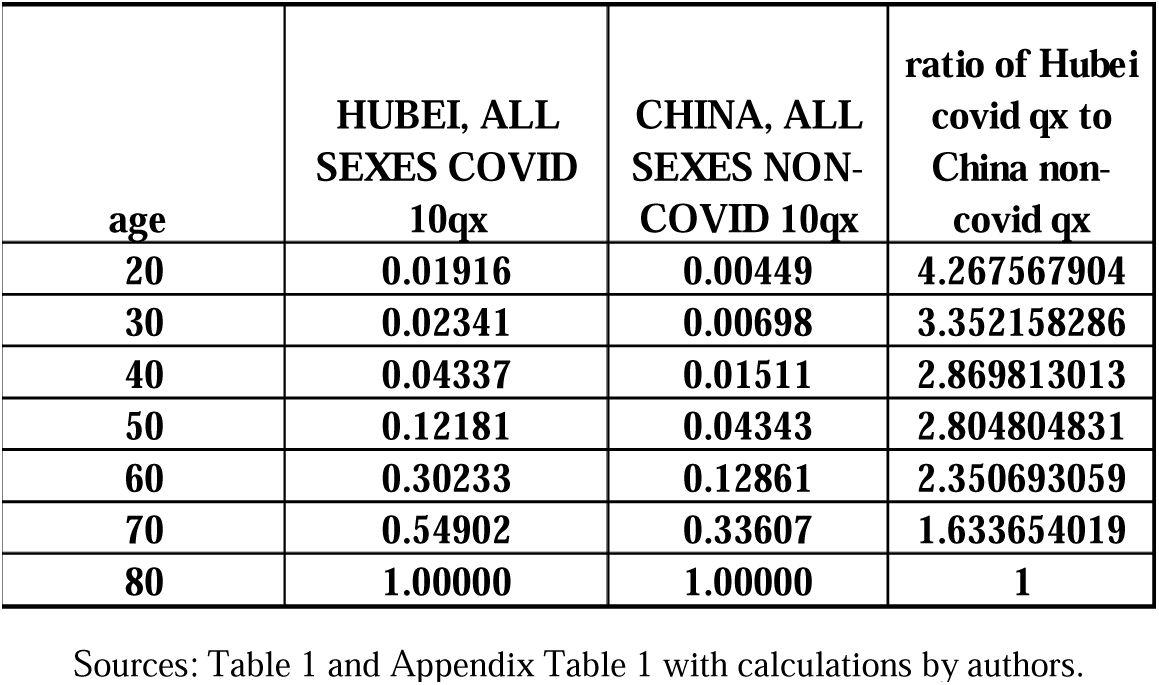
Ratio of COVID-19 _n_Q_x_ to Non-COVID-19 _n_Qx.

**Figure 1.**
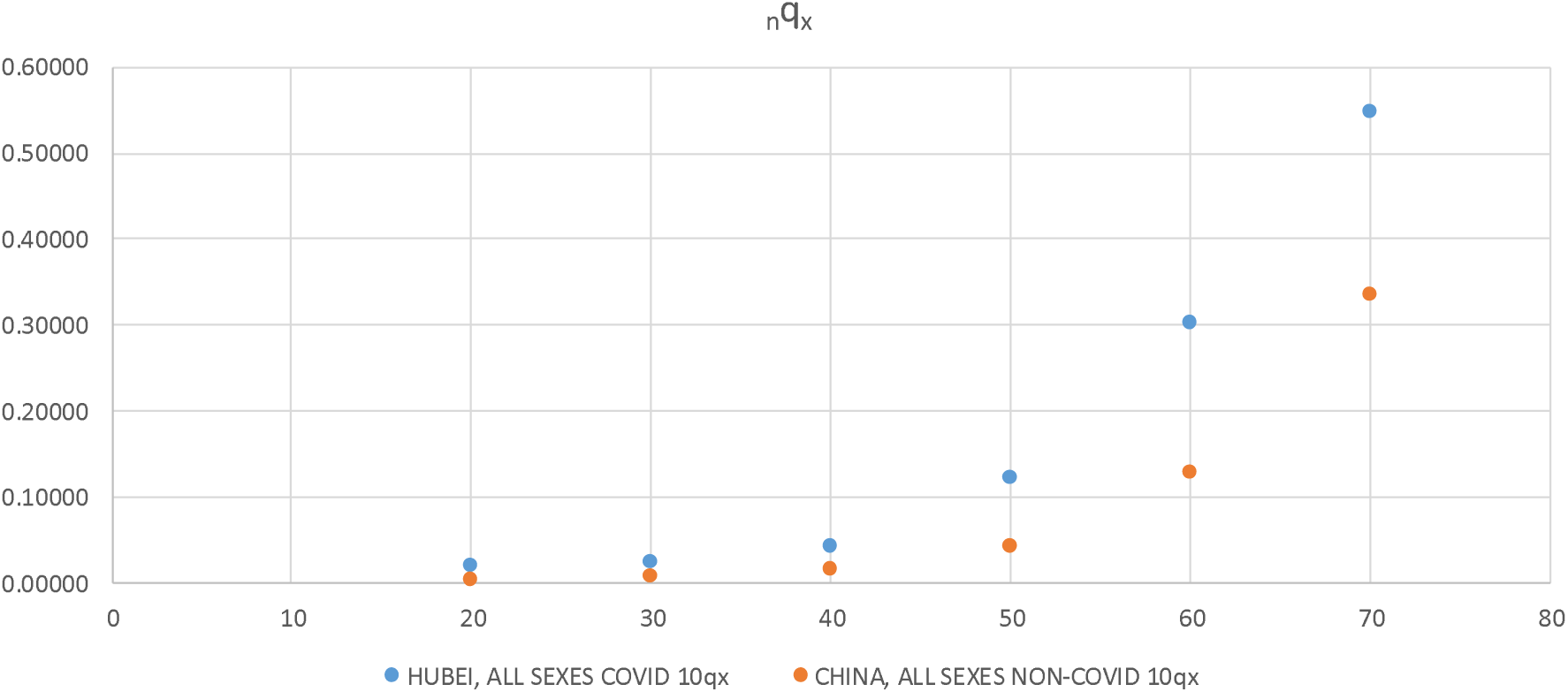
All sexes, Hubei 2020 Covid-19 Victim _n_q_x_ vs. China 2016 non-Covid

**Figure 2.**
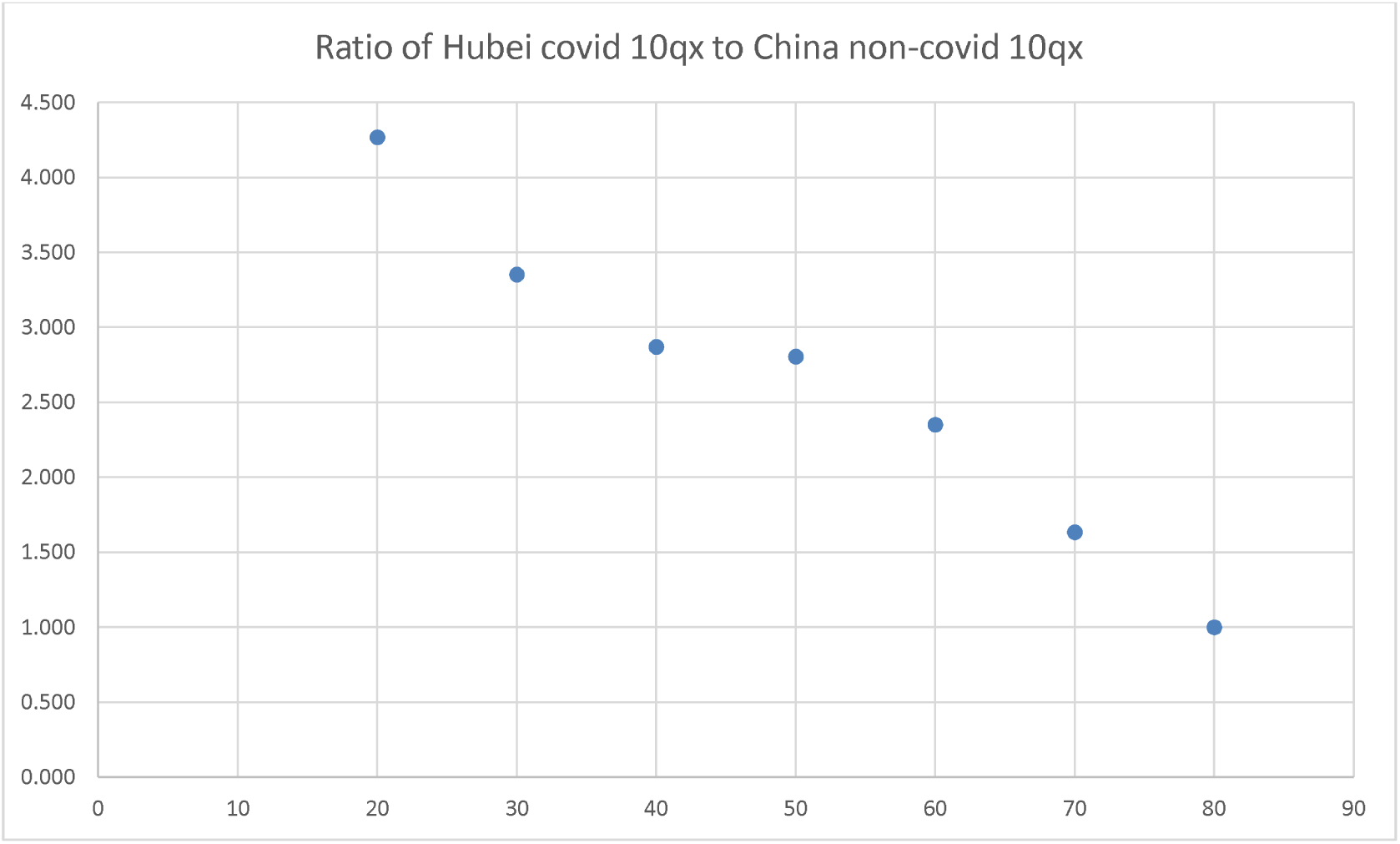
Ratio of _10_q_x_ values for the Hubei COVID-19 victims in 2020 and the _10_q_x_ values derived from the 2016 life table for China.

At age 20, the probability of a person with Covid-19 dying from it (and no other causes) in Hubei before reaching age 30 is 4.27 times higher than the probability of a person aged 20 in China dying before reaching age 30 of any other cause other than Covid-19, which is 0.00449; at age 30, the probability is 3.35 times higher, age 40 it is 2.87 times higher, age 50, 2.80 times higher; and at age 60 the probability of a person with Covid-19 dying of it before reaching age 70 in Hubei is 1.63 times higher than the probability of a person age 70 in China dying of any cause other than Covid-19 before reaching age 80, which is 0.33607.

Given that the life table for China as a whole applies to Hubei Province, Table 2 shows the increased mortality risk generated by COVID-19 on Hubei’s population. Looking in the other direction, Table 2 shows the increased mortality risk generated by COVID-19 on China’s population given that the Hubei COVID-19 life table applies to China as a whole.

## Discussion

Table 2 shows that those aged 20 in Hubei Province who contracted COVID-19 were at a higher probability of dying from it than were non-infected people in China aged 20 dying from all other non-COVID-19 causes. This probability increases monotonically with age, but declines relative to the probability for those in China not exposed to COVID-19. As such, beyond age 50 it appears to be more deadly for them than was smallpox in the 18^th^ and 19^th^ centuries when Bernoulli presented a strong case that among those who are infected, about 12 percent of them will die in any given age group (Gehrman, 2014). Another way to look at the risk of dying is to construct a survival model. Using the data in Table 1 for the population aged 20 and over, we constructed a Gompertz model from the deaths found in column 7 using the survival analysis procedure found in the NCSS software package, release 12 (NCSS 2020). Exhibit 1 provides the details of the results.

(EXHIBIT 1 ABOUT HERE)

With a coefficient of determination equal to 0.95 and a slope parameter of 0.1273, we are presented with a well-fit model in which the “actuarial ageing rate” is very high - the rate of dying increases very rapidly with age (Kirkwood, 2014: 1). These comparisons and the ratios found in the last column of Table 2 are telling: Once infected, there is a high probability of dying that increases exponentially with age.

The Hubei data are not a random sample, which usually would mean that classical inferential statistics such as confidence intervals and margins of error are not appropriate. However, we can take the “superpopulation” perspective (Deming, 1953; Godambe and Thompson, 1986. Swanson and Beck, 1994; Swanson and Tayman, 2012: 171-173; Swanson, Tayman, and Beck 1995), which provides an alternative framework for inference, an approach often found in wildlife studies, where scientific sampling is extremely difficult (e.g., Durkin and Cohen, 2019; Wen et al., 2011). While not perfect, this perspective does provide some idea of the uncertainty affecting the Hubei _10_q_x_ values. With the superpopulation perspective in mind, we use an approach developed by Chiang (1960) to generate uncertainty measures. Starting with the unbiased estimate of probability of death q^ = D/ N, where the number of deaths is denoted by D within a given age interval and N is the number at the beginning of the interval, Chiang (196)) showed that the standard deviation can be estimated as sqrt(q^*(1- q^))*N, which leads to the standard error (se) of death in the interval in question of sqrt(q^*(1- q^)/N). Translated into the terms we use in Table 1 and elsewhere in this paper, we have se = sqrt(_10_q_x_*(1-_10_q_x_)/k), where k = number alive at age x. Table 3 provides the standard errors and 95% confidence intervals for the _10_q_x_ values of the Hubei COVID-19 victims (Table 1). For age zero, there are no reported deaths so a confidence interval is a moot point as is the case for age 80 and over where the probability of dying is 1.00. For the other age groups, the estimated standard errors range from 0.00173 for age group 30-39 to 0.00795 for age group 70-79. Even where the standard error is the highest, the 95% confidence interval for age group 70-79 is relatively narrow, from 0.530 to 0.565. If nothing else, these results show the impact that a large sample can have in the reduction of statistical uncertainty.

**TABLE 3.** Confidence Intervals for the Hubei _10_q_x_ values in Table 1.

| **Age Interval** | **$_{10}q_x$** | **Standard Error** | **95% lower limit** | **95% upper limit** |
| --- | --- | --- | --- | --- |
| **0-9** | **0.00000** | **NA** | **N/A** | **N/A** |
| **10-19** | **0.01805** | **0.00568** | **0.0069134** | **0.029186676** |
| **20-29** | **0.01916** | **0.00228** | **0.0146905** | **0.023622507** |
| **30-39** | **0.02341** | **0.00173** | **0.0200068** | **0.026805083** |
| **40-49** | **0.04337** | **0.00220** | **0.039055** | **0.047679238** |
| **50-59** | **0.12181** | **0.00327** | **0.1154053** | **0.128221326** |
| **60-69** | **0.30233** | **0.00496** | **0.2926171** | **0.312049787** |
| **70-79** | **0.54902** | **0.00795** | **0.5334355** | **0.564597614** |
| **80+** | **1.00000** | **N/A** | **1** | **1** |

Source: Table 1 with calculations by authors

## Concluding Remarks

An immediate question is how well the Hubei _10_q_x_ values apply elsewhere. As a starting point, we note that Verity et al. (2020: 669) found that the case fatality ratios they estimated by age from the Hubei data were consistent with those international cases stratified by age. This consistency suggests that the COVID-19 life table (Table 1) may apply beyond Hubei Province. Another consistency is that the conditional probability of dying from COVID-19 from age 20 onward increases monotonically for those who are infected, which follows a general age-related mortality pattern (Kintner, 2004; Yusuf, Martins, and Swanson, 2014). Relative to the probability of dying in China from all other causes for those not infected, however, it declines with age, as is seen in the last column of Table 2. This decline is a result of the age-related increase in the probability of dying from one or more of a number of competing causes, which, in the case at hand is manifested in the fact that non-COVID-19 deaths in China among the uninfected were generally increasing at a faster age-related rate than were the COVID-19 deaths to the infected in Hubei.

If one is interested in comparing Hubei to a country other than China, one should start by comparing the _10_q_x_ values for China shown in Table 2 with those for the country in question. If they have similar _10_q_x_ values, the overall pattern will hold in that there will be a decline in the ratio in going from 20 to age 70 (for the reasons just stated), but it may not be as steep as the decline in the ratio seen for China, which would be the case with a country that has higher _10_q_x_ values than China at the younger ages and lower ones at the older ages. The pattern also may not be monotonic, which would be the case for a country that had the same _10_q_x_ values as China at every age except 50, where it was substantially lower, which would result in a “bump” in the trend at age 50.

As noted by Verity et al. (2020) and Tian et al, (202) initial analysis of the survival data of Wuhan needs revision after the epidemic unfolds. In the meantime, we trust that the results presented here will assist hospital and clinical settings in long-term and current management.

## Data Availability

All found in the paper or at publicly available online sources

## Appendix

Let *l (x, t)* be the infected individual at age *x* and at time *t*. The probability of survival of infected who will live for *k* time units since acquiring the infection at *t* say, _*k*__*p(x, t)*_, is computed as

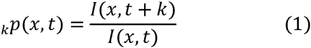

and probability of survival for all infected at age *x* to survival until *k* is

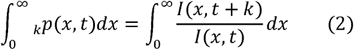

The probability of survival for all infected during (*t, t* + *k*) will be

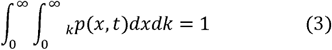

Then, the probability of dying of an individual who is at age *x* to die within (*t, t* + *k*) will be

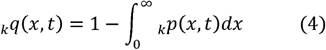

The decadal probability of dying from (4) would be

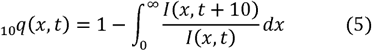

## Acknowledgments

None

---

**Exhibit 1.**
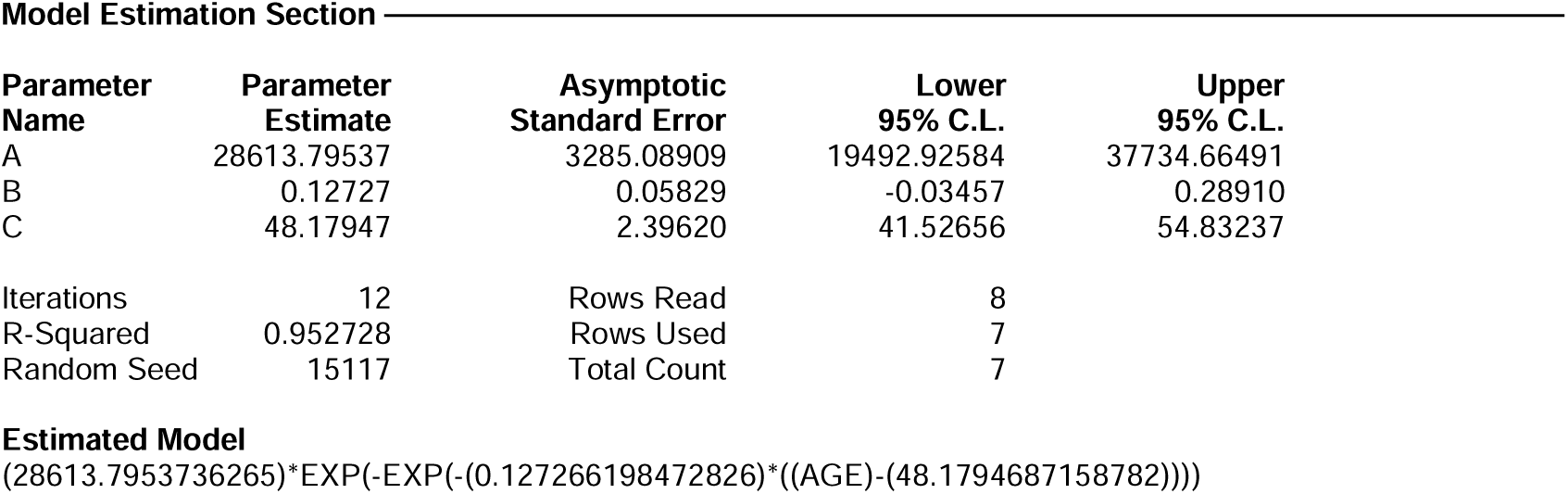
NCSS Gompertz Model Results

**Appendix Table 1.**
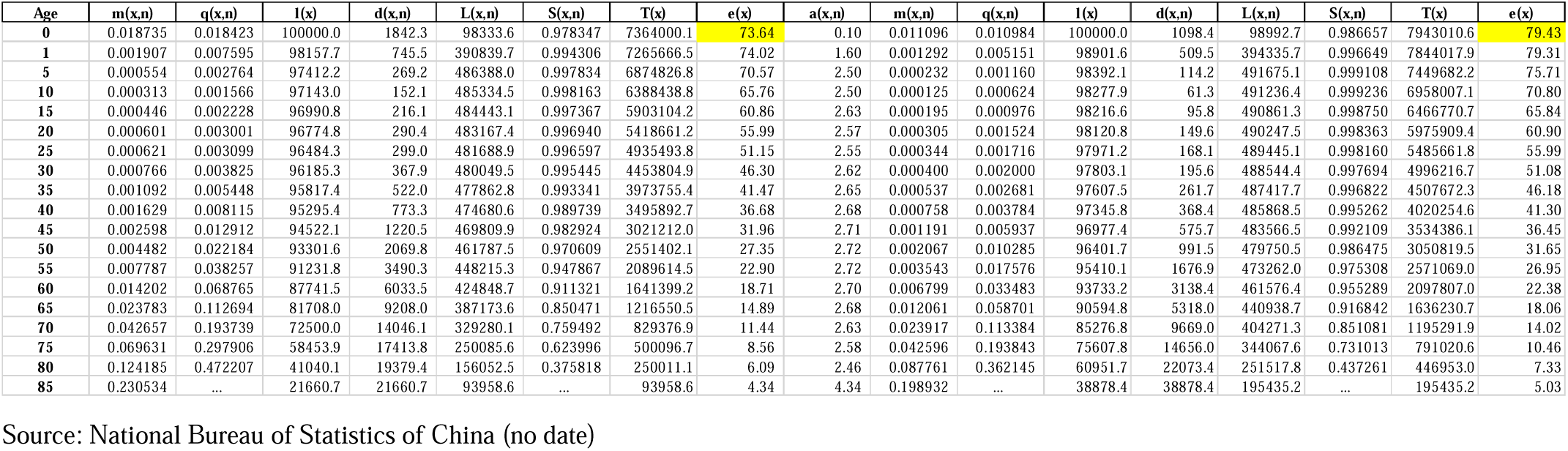
Abridged life table by Gender, China 2016

## References

Andersen, P., V. Canuas-Romo, and N. Keiding, (2013). Cause-specific measures of life years lost. Demographic Research 29: 1127–1152. DOI: 10.4054/DemRes.2013.29.41

Andrasfay T., and N. Goldman (2021). Association of the COVID-19 Pandemic with Estimated Life Expectancy by Race/Ethnicity in the United States. JAMA Netw Open. 4(6):e2114520. doi:10.1001/jamanetworkopen.2021.14520.

Britannica (no date). Hubei (https://www.britannica.com/place/Hubei).

Castro, M., S. Gurzenda, C. Turra, T. Andrasfay, and N. Goldman (2021). Reduction in life expectancy in Brazil after COVID-19. Nature Medicine 27, 1629–1635 https://doi.org/10.1038/s41591-021-01437-z

Chan E., D. Cheng, and J. Martin. (2021) Impact of COVID-19 on excess mortality, life expectancy, and years of life lost in the United States. PLoS ONE 16(9): e0256835. https://doi.org/10.1371/journal.pone.0256835.

Chen N., et al. (2020) Epidemiological and clinical characteristics of 99 cases of 2019 novel coronavirus pneumonia in Wuhan, China: a descriptive study. Lancet 395: 507–13.

Chiang, C. (1960). A stochastic study of the life table and its Applications II. Sample Variance of the observed expectation of life and other biometric functions. Human Biology 32 (3): 21–238.

Durkin, M., and J. Cohen. (2019). Estimating Avian Road Mortality When Only a Single Observer Is Available. The Journal of Wildlife Management 83 (1): 100–108.

Fergany, N. (1971). On the Human Survivorship Function and Life Table Construction. Demography 8 (3): 331–334.

Gehrman, R. (2014). The impact of smallpox and vaccination in Northern Germany in the 18th and 19th centuries. Przeszlość Demograficzna Polski 36: 39–54.

Godambe, V. and M. Thompson (1986). Parameters of Superpopulation and Survey Population: Their Relationships and Estimation International Statistical Review 54 (2): 127–138.

Goldstein J., and R. Lee (2020. Demographic perspectives on the mortality of COVID-19 and other epidemics. Proceedings of the National Academy of Sciences of the United States of America.117(36): 22035–22041. pmid:32820077.

Heuveline P. (2021) The Mean Unfulfilled Lifespan (MUL): A new indicator of the impact of mortality shocks on the individual lifespan, with application to mortality reversals induced by COVID-19. PLoS ONE 16(7): e0254925. https://doi.org/10.1371/journal.pone.0254925.

Huang, C., et al. (2020). Clinical features of patients infected with 2019 novel coronavirus in Wuhan, China. Lancet 395: 497–506.

Islam, N., D. Jdanov, V. Shkolnikov, K. Khunti, I. Kawachi, S. Lewington, and B. Lacey. (2021). Effects of covid-19 pandemic on life expectancy and premature mortality in 2020: time series analysis in 37 countries. BMJ 375 doi: https://doi.org/10.1136/bmj-2021-066768.

Kintner, H. (2004). The Life Table, pp. 301–340 in J. Siegel and D. Swanson (eds). The Methods and Materials of Demography, 2^nd^ Edition. Academic/Elsevier Press: Los Angeles.

Kirkwood, T. (2014). Deciphering death: a commentary on Gompertz (1825) ‘On the nature of the function expressive of the law of human mortality, and on a new mode of determining the value of life contingencies. Philosophical Transactions of the Royal Society B 370 (http://dx.doi.org/10.1098/rstb.2014.0379).

National Bureau of Statistics of China (no date). 2016 Life Table of China by Sex.

NCSS 2020 Statistical Software (2020). NCSS, LLC. Kaysville, Utah. (https://www.ncss.com/software/ncss/)

Pifarréi Arolas, H. E. Acosta, G. López-Casasnovas, A. Lo, C. Nicodemo, T. Riffe and M. Myrskylä. (2021). Years of life lost to COVID-19 in 81 countries. Scientific Reports 11: 3504 https://doi.org/10.1038/s41598-021-83040-3.

Ritchie, H., E. Mathieu, L. Rodés-Guirao, C. Appel, C. Giattino, E, Ortiz-Ospina, J. Hasell, B. Macdonald, D. Beltekian and M. Roser. (2022). Coronavirus Pandemic (COVID-19). Coronavirus Pandemic (COVID-19) – the data - Statistics and Research - Our World in Data (accessed January 19th, 2022).

Statistica (no date). Population distribution in China in 2020, by five-year age group and gender. (https://www.statista.com/statistics/1244036/population-distribution-by-five-year-age-group-and-gender-in-china/).

Stroup, A, H. Cho, S. Scoppa, H. Weir, and A. Mariotto. (2014). The Impact of State-Specific Life Tables on Relative Survival. Journal of the National Cancer Institute, Monographs 49:218–227.

Swanson, D. (2021). An Example of Converting Clinical Study Mortality Data into a Life Table: The U.S. Population with Sickle Cell Disease. Open Journal of Public Health. 3 (1): 1–5.

Swanson, D. and D. Beck. (1994). A New Short-term County Population Projection Method”. Journal of Economic and Social Measurement 20: 25–50.

Swanson, D., and J. Tayman. (2011). Subnational Population Estimates. Springer B.V. Press. Dordrecht, Heidelberg, London, and New York.

Swanson, D. and L. Tedrow (2018). A Bio-demographic Perspective on Inequality and Life Expectancy: An Analysis of 159 Countries for the Periods 1970-90 and 1990-2010. pp. 577–613 in C.R. Rao and A. Rao (eds.), Handbook of Statistics, Vol. 38. Elsevier Press.

Swanson, D. S. Chow, and T. Bryan (2020), Constructing Life Tables from the Kaiser Permanente Smoking Study and Applying the Results to the Population of the United States.” pp.115–152 in B. Jivetti and M. N. Hoque (eds.). Population Change and Public Policy. Springer B.V. Press. Dordrecht, Heidelberg, London, and New York.

Swanson, D., D. Beck, and J. Tayman (1995) “On the Utility of Lagged Ratio-Correlation as a Short-term County Population Estimation Method: A Case Study of Washington State.” Journal of Economic and Social Measurement 21:1–16.

Swanson, D., P. Morrison, D. Poston, S. Krantz, and A. Rao. (2022). America’s Post-pandemic Future: A Demographic Perspective. PAA Affairs (https://www.populationassociation.org/browse/blogsãCommunityKey=a7bf5d77-d09b-4907-9e17-468af4bdf4a6).

Tian, R., et al. (2020). Clinical characteristics and survival analysis in critical and non-critical patients with COVID-19 in Wuhan, China: a single-center retrospective case control study. Scientific Reports 10 17524. https://doi.org/10.1038/s41598-020-74465-3

Trias-Llimós S, and U. Bilal. (2020). Impact of the COVID-19 pandemic on life expectancy in Madrid (Spain), Journal of Public Health 42 (3): 635–636, https://doi.org/10.1093/pubmed/fdaa087.

Venkataramani A., R. O’Brien, A. Tsai (2021). Declining Life Expectancy in the United States: The Need for Social Policy as Health Policy. JAMA 325 (7):621–622. doi:10.1001/jama.2020.26339.

Verity, R., et al., (2020). Estimates of the severity of coronavirus disease 2019: A model-based analysis. Lancet 20 (6): 669–677.

Wang, H., L. Dwyer-Lindgren, K. Lofgren, J. Rajaratnam, J. Marcus. A. Levin-Rector, C. Levitz, A. Lopez, and C. Murray (2012). Age-specific and sex-specific mortality in 187 countries, 1970–2010: a systematic analysis for the Global Burden of Disease Study 2010. The Lancet 380 (9859): 2071–2094.

Wen, Z., K. Pollock, J. Nichols, and P. Waser (2011). Augmenting Superpopulation Capture—Recapture Models with Population Assignment Data. Biometrics 67 (3): 691–700

Woolf, S. (2021). Effect of the covid-19 pandemic in 2020 on life expectancy across populations in the USA and other high income countries: simulations of provisional mortality data. BMJ 373 doi: https://doi.org/10.1136/bmj.n1343.

Yusuf, F, J. Martins, and D. Swanson. (2014). Multiple Decrement Life Tables, pp. 215–299 in Methods of Demographic Analysis. Springer B.V. Press. Dordrecht, Heidelberg, London, and New York.

